# Predicting Whom to Test is More Important Than More Tests - Modeling the Impact of Testing on the Spread of COVID-19 Virus By True Positive Rate Estimation

**DOI:** 10.1101/2020.04.01.20050393

**Authors:** Paul Rodriguez

## Abstract

I estimate plausible true positive (TP) rates for the number of COVID-19 tests per day, most relevant when the number of test is on the same order of magnitude as number of infected persons. I then modify a standard SEIR model to model current growth patterns and detection rates in South Korea and New York state. Although reducing transmission rates have the largest impact, increasing TP rates by ∼10% in New York can have an impact equal to adding tens of thousands of new tests per day. Increasing both TP rates and tests per day together can have significant impacts and likely be more easily sustained than social distancing restrictions. Systematic and standardized data collection, even beyond contact tracking, should be ongoing and quickly made available for research teams to maximize the efficacy of testing.

## Introduction

As the COVID-19 virus spreads there are wide calls for increased testing to identify cases and slow the spread. Colburn (2020) has pointed out that we need to model the effects of testing, contact tracking, isolation, and related interventions in order to better use resources. Currently, (March 28^th^, 2020), South Korea seems to have successfully contained the spread using a variety of strategies, including wide-spread testing and tracking. However, unless the number of tests approaches the size of the population, or the size of all possibly infected persons, there will be some criteria for choosing whom to test. Choosing whom to test is a decision problem for which predicting who is pre-symptomatic would be most beneficial. How beneficial depends on the true positive rate (i.e. ratio of true positives to sum of true positives and false negatives). But the difficulty in modeling the true positive (TP) rate of testing and related strategies is that the number of false negatives (misses) are unknown.

Here, using preliminary online data reports, I derive rough approximations of TP rates from numbers of tests relative to a population to produce a ROC (Receiver Operating Characteristic) curve, which, to my knowledge, has not been presented yet even in such a rough form. I then modify a standard SEIR model (R, Epidynamics package) to use TP rate to decrease the pool of infected individuals. Using model parameters published elsewhere, a grid search of TP rates and beta (time to contact) values shows that, in regions of parameter space that match empirical data, increasing TP rate can have critical impacts in slowing spread of the virus. TP rate can increase with large increases in testing or by improving the prediction on whom to test for. In fact, public health agencies should be gathering data for such endeavor which could be done independently of any need for material resources of testing kits.

## Methods

### Estimating TP Rate

It is not known how effective limited COVID-19 testing can be for slowing the spread of the virus, but there is some preliminary cases that can help give rough estimates of TP and FP rates from the number of testing per day. For example, South Korea health board reported to have been testing up to about 13000-15000 individuals per day, from when the maximum increase in cases was near 1000 per day at the end of February up to the end of March (South Korean Public Health Board). The new cases rate has dropped to about 100 per day (March 28th). The ratio of 15-to-1 ratio of tests-per-new-cases at the peak is much higher than in US states like New York and California currently, and these and others states are trying to rapidly increase these number (COVID Tracking Project). Studies suggest that COVID-19 can incubate for an average of 6.4 days (e.g. Prem et al., 2020). In the standard SEIR epidemiological model new infected individuals (*I*) come from the exposed incubating group (*E*). Therefore, if one assumes that the COVID-19 test is near 100 percent accurate for candidates from both groups, and that results are produced in 1 day, then at the peak of new cases in South Korea there were about 6000 cases in group *E* and the TP rate was ∼ 0.16. Of course, this is an approximation that does not take into account many other factors, such as travelers, individuals who have symptoms but do not have COVID-19, retesting, or the accuracy of the test itself, as opposed the accuracy of selecting whom to test.

Another example to consider is the Diamond Princess cruise ship case. According to the CDC all individuals were tested over several weeks, some multiple times (Moriarty, et al., 2020). There were ∼10% of individuals on board who tested positive and showed symptoms, ∼10% who tested positive and showed no symptoms at the time of testing, and 80% negative. This suggests that if you have a number of tests on the order of the number of infected individuals you would only get about 50% TP rate. If you had more tests and only considered symptoms as a predictor, then after the first 10% of the population (with symptoms), the TP rate would increase at the baseline rate of 1 out 9 tests. Contact tracking could help, but in some sense the whole ship was in the sphere of possible contact.

Using these two examples one could produce a plausible ROC curve of sensitivity (TP rate) vs specificity (FP rate), Figure 1. In this case, the values on the axis are percentages from 0 to 1. Importantly, the graph is not easily parameterized by the absolute number of tests but should be thought of as parameterized by relative values as follows. If we think of the cruise ship example, we imagine that all the passengers & crew are given a score that predicts their likelihood of having COVID-19, and then tested in sequence of rank 1 to rank P, the population size, according to the score. As we test from 1/P to P/P fraction then the TP rate and FP rate will go from 0 to 1 as function of time. In other words, as we test more and more of a population, the TP rate will increase as shown.

**Figure 1.**
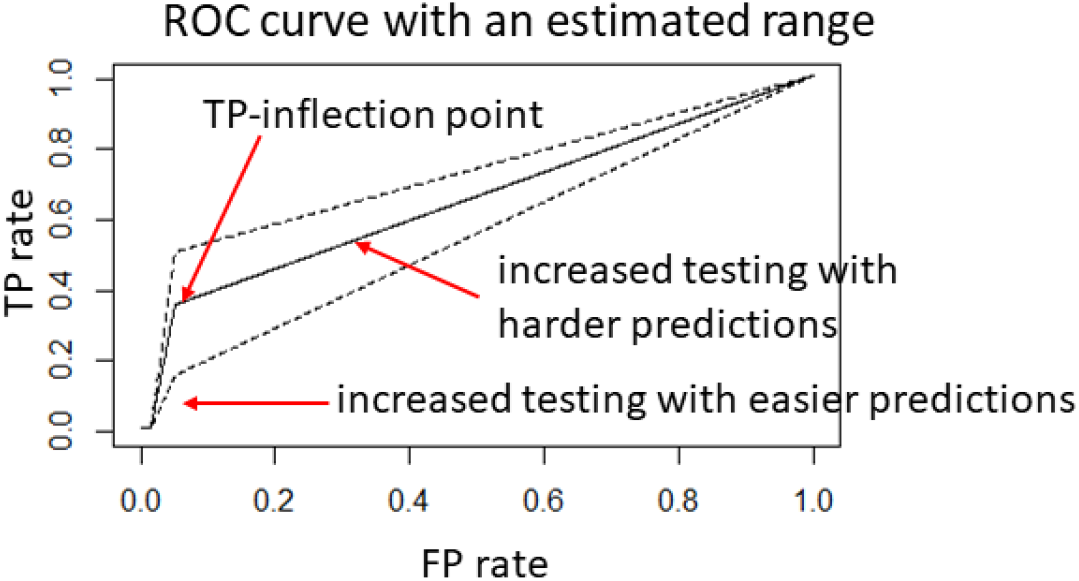
An estimated ROC curve of TP rate and FP rate, with an inflection point given by a parameter called TP-infpt, i.e. the TP rate when the number of tests per day is maximal but still somewhat lower than number of persons thought to be possibly infected. The region between the dashed lines are suggested plausible regions of rate values based on reported data. In simulations for South Korea and New York state the initial TP rate at the origin is 0.01 (1%) and rises quickly. The time before it rises and hits the inflection point depends on number of days and speed with which health agencies ramped up their testing capacity.

Conversely, we can also imagine the curve is parameterized as a function of the score threshold. As we get more and more tests our threshold for who gets tested will decrease, and TP rate will increase. Of course, the false positive rate (i.e. individuals who are predicted to be positive but are negative) will increase as well but that cost is much less than misses (false negatives). A third way to consider the parameterization is a function of number of infected individuals (*I*), which is perhaps most relevant for current situation. If our testing is on the order of magnitude as *I*, and if we think we need to have 15* *I* tests, then the units could be considered multiples of *I*. In either case, the shape of the graph is the same but the increase in the efficacy of testing will depend on whether we can increase the number of tests, or decrease the pool of potentially infected persons, or just test for more days. If we could also improve the score thresholding, then that could increase the whole shape of the graph and change the inflection point.

In the next sections the SEIR model is adapted to incorporate TP rates, and using parameters published elsewhere, the model is applied to South Korea and New York state. A vital question is how much impact an increase in testing will have in New York. We run the model to show tradeoffs and potential benefits of more testing, better predictions, and possible growth curves.

### Modifying the SEIR Model

The SEIR infectious disease spread model was implemented in the R package EpiDynamics (Baquero & Marques, 2015), using methods from Keeling & Matt (2008). I modified the model so that the change in number of persons infected, *dI/dt*, is decreased by [*-1 * TP rate * I*] and the recovered pool (R) is increased by the same amount, which implies detected cases are immediately removed from the susceptible pool (S). The TP rate was given by the curve in Figure 1 with 4 parameters; TP-infpt, is the TP rate inflection point, t1=‘time-to-start’ is the time during which few tests were done as the epidemic started, t2=‘time-to-ramp-up’ is the time at which maximum daily tests were given, and P for population size. For example, South Korea did not start with 15000 tests but took some days to get organized and ramp up. Based upon reports, tests increased rapidly and steadily after February 10th up to March 28th (South Korea CDC). Use a starting time around mid-January, I set t1=20, t2=68. The other parameters are set as described in Table 1. Note that beta was varied to model and test different effects of TP rates, and mu was set to 0 to ignore birth/death rates.

**Table 1.**
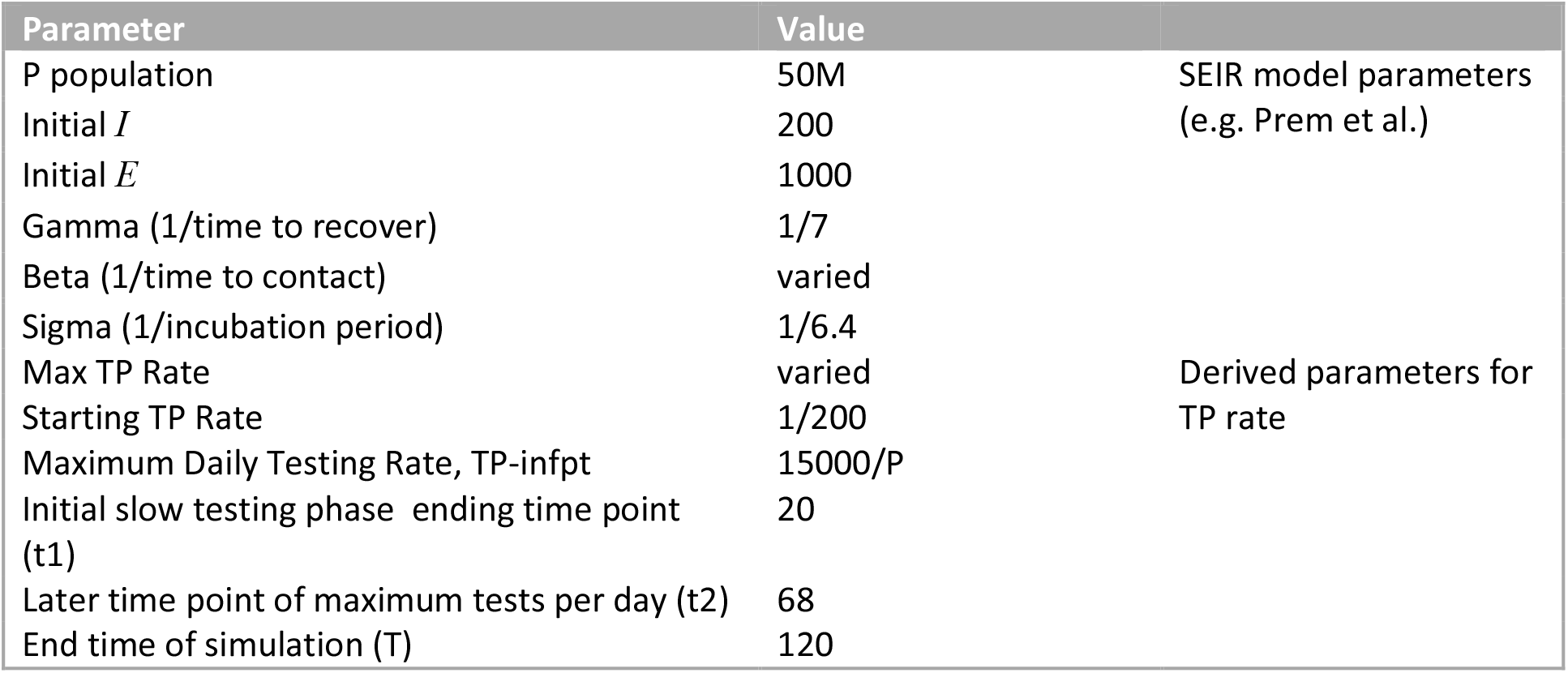
Simulation parameters for South Korea simulation. SEIR model parameters were taken from Prem et al., and other parameters are derived from analysis here for TP rate.

I ran the model for TP-infpt = 0.1 to 0.5, beta= 0.22 to 0.42 (R0∼beta*7, or R0∼1.5 to 3.0, where 1/7 is recovery rate, and beta is the contact rate). The contour plot in Figure 2 shows the maximum values reached in simulations (up to t = 120). The goal of the simulations is not to precisely match reported number of infected persons, which likely do not reflect true values, but to match the general time of peaking in the growth curve to within an order of magnitude. The growth plot in Figure 2 shows an example where the TP-infpt and beta parameters match the empirical growth curve for South Korea (e.g. www.worldodemeters.info). For South Korea, the peak value of infected cases was about 7300, and peak days were about 45-60 days (early to mid-March). The effective TP rate can have a big impact on whether growth is suppressed or increases because it essentially counteracts R0, the reproduction number (i.e. transmission rate). However, when R0 is very high (i.e. beta > 0.40) the impact of TP rate is relatively smaller, which is why Figure 2 has non-linear contour lines in that region. Even so, as the example growth plot shows, the TP rate can make a big difference over time.

**Figure 2.**
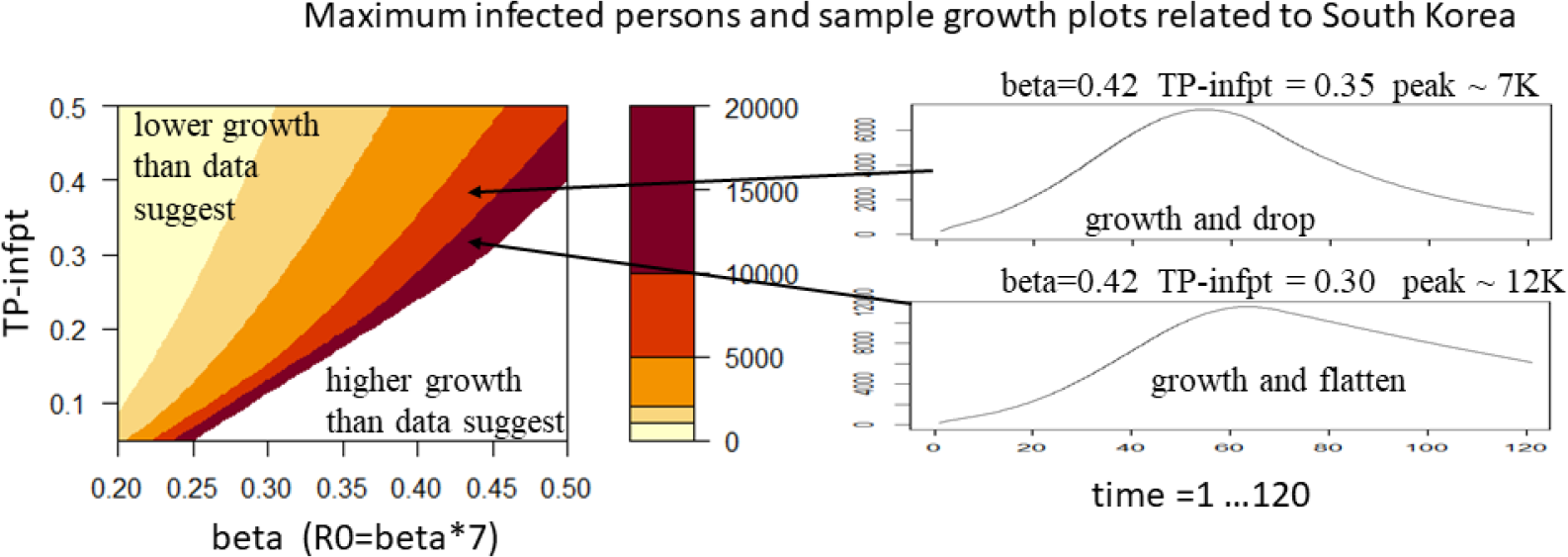
The general regions of growth curves for model parameters shows how TP rates and beta tradeoff for the case of South Korea. Sample disease spread growth curves are shown for where number and shape of growth are similar, within an order of magnitude, to South Korea, particularly the curve that drops after peaking.

I also ran the model to compare to New York state. Based on reported testing numbers (COVID Tracking Project) the early part of March had wide variety and missing numbers. But after March 12 testing increased steadily. Using a rough starting period around February 1st, I set t1=42. I set t2=58 and maximum daily test rate to 25000 because around 23000 tests per day were conducted March 28^th^. Other parameters are same as South Korea, except population, and are described in Table 2.

**Table 2.**
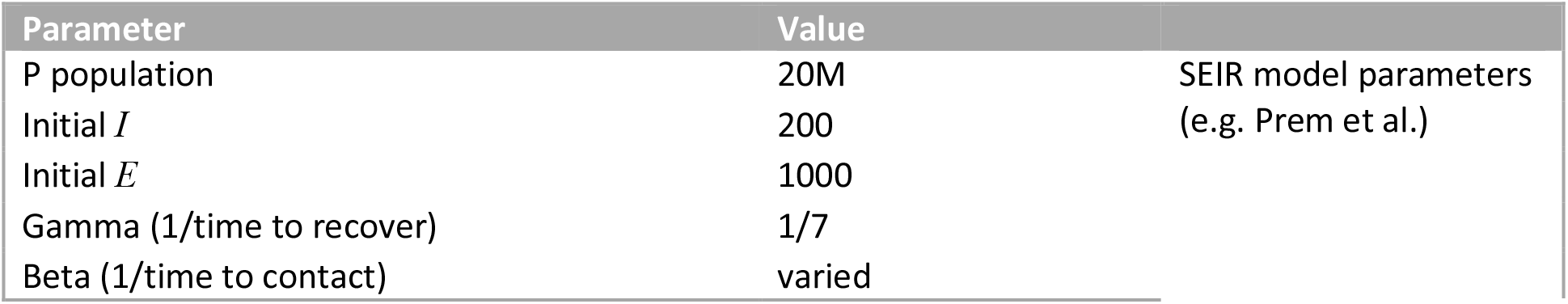
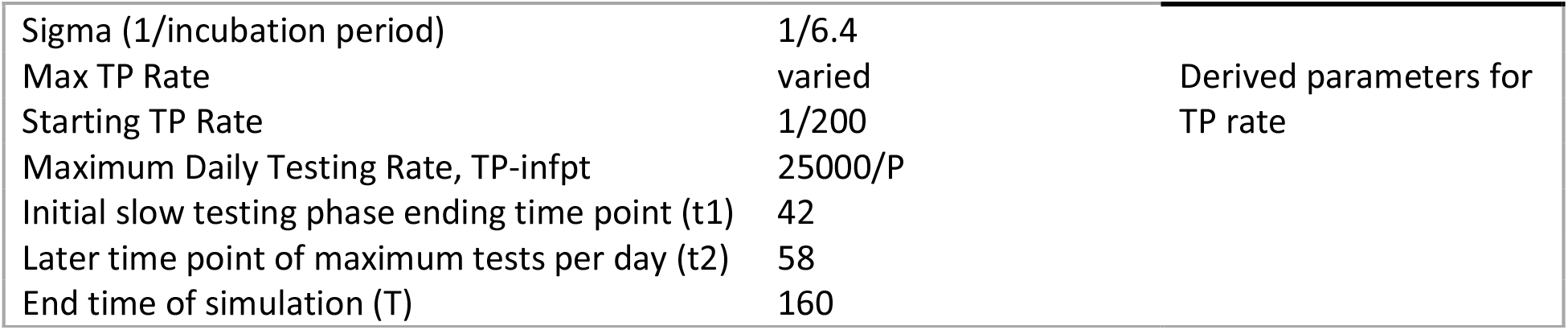
Simulation parameters for New York state simulation. Notice the testing phase time points are higher but steeper than South Korea.

The contour plot in Figure 3 shows maximum values reached during the simulation, and a sample growth curve with a peak value of 188K infected persons, and a value on day 58 near the currently reported number (∼50K). The true number is likely higher, but it gives a starting point to consider tradeoffs in parameters. The parameters for the sample growth curve were beta =0.42 (same as Figure 2) and TP-infpt 0.15. It is reasonable that TP-infpt is lower for New York than South Korea based on reports that South Korea has been more aggressive in testing and tracking contacts.

**Figure 3.**
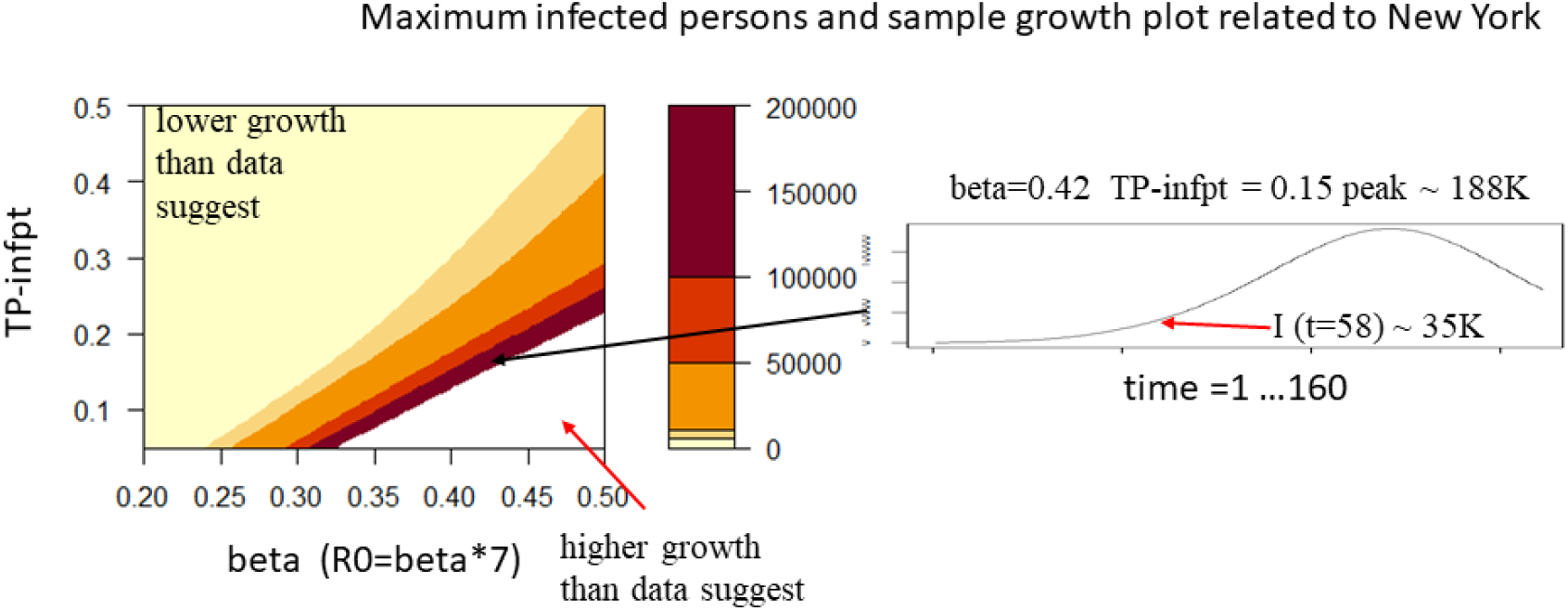
The general regions of peak growth for model parameters shows how TP rates and beta trade-off for the case of New York state. A sample growth curve is shown for the region of parameter values where number of infected person are the same to the current count around day 58, to within an order of magnitude.

Using the model values at day 58 as initial values, I ran the model again from that time point but with slight variations. It turns out that increasing TP-infpt rate by 10%, from 0.15 to 0.165 after day 58 has the same effect as increasing the number of tests by 50% to 37500 tests per day after day 58. The effect in both cases is to cut the number of cases at the peak by about 25% from 188K about day 120 to about 140K on or about day 108. These values should not be taken as predictions about absolute infected counts that might occur, but rather they suggest the relative importance of improving the efficacy of testing and increasing the number of tests.

Figure 4 extends the simulation for a range of new parameter values that take effect after day 58. In the contour plot, starting from the lower left corner with TP-infpt rate of 0.15 and test per day at 25K, increasing TP rates have a bigger impact on decreasing the peak value of infected person. In the sample growth curve plot, change both TP-infpt rate and number of tests per day by 50% will dramatically change the growth curve.

**Figure 4.**
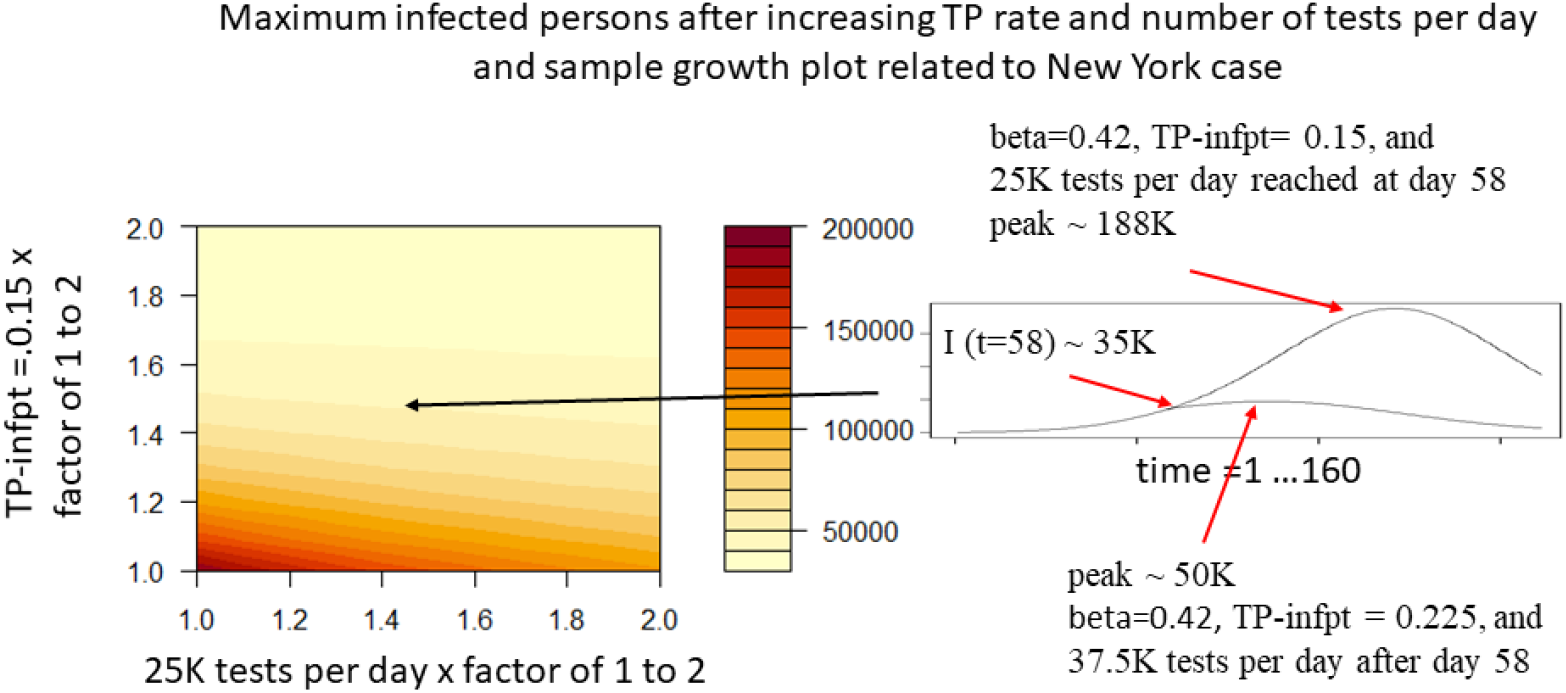
Simulating the effects of increasing TP rates and maximum number of tests per day. The contour plot shows maximum number of infected persons as TP-infpt rate and tests per day are increased. The increases are relative to the values using in the growth plot in Figure 3 which is similar to data observed in New York. The sample growth plot suggests that increasing TP rates and tests by a factor of 1.5 will start flattening the growth curve immediately.

## Discussion

I presented a rough estimate of a ROC curve to simulate how increasing the number of tests slow the spread of COVID-19. An important point that is easily overlooked is that knowing the TP rate relative the number infection cases is likely more useful than knowing the absolute number of tests or number of tests per capita, as is often reported in the news media. In fact, the relation between the number of tests and proportion of detected cases out of all positive cases is indicated by the TP rate. In the current crises, this is unknown. Nonetheless, with plausible values of the TP rate one can model the impact of increasing TP rates. With a modified SEIR model, I have shown that simulations of virus spread can produce valid estimates of currently infected persons and enable estimates of future growth under different scenarios.

It has become painfully obvious that the number of COVID-19 cases in New York will still be increasing for some time. A critical strategy is to reduce the reproduction number by reducing contact and movement because the growth factor has an exponential effect. However, at this point in time it may not by itself slow the spread enough unless restrictions become draconian and long lasting. More testing and quarantining will help mitigate the rise, but if testing is going to copy the South Korean numbers, the testing in New York must increase faster than the virus spread, perhaps to get up to 15 to 1 ratio of new cases (as of March 28th, that would be about ∼75000 tests per day). But just as important is to increase efficacy by collecting good data to better score and allocate tests. A good scoring model and allocation strategy, that increases TP rate by a one or two percentage points could have the same impact as increasing the number of tests by tens of thousands. The scoring model perhaps could include contacts and locations, along with demographics, weather, type of places visited, time of day, and anything else that could help build a robust model on top of virus spread models and tracking contacts. In some high profile machine learning competitions, intensive application and modeling efforts have shown improvements of 10 percentage points in accuracy (e.g. Netflix Prize, Wikipedia, Zillow Landing Pages). There should be low technical or material barriers to gathering and using data, and I would argue that health agencies should start doing so and making the anonymized data available.

## Data Availability

Data in the manuscript is publicly available at the URLs given in the references.

